# Altered markers of mitochondrial function in adults with autism spectrum disorder

**DOI:** 10.1101/2023.06.26.23291908

**Authors:** Kathrin Nickel, Mia Menke, Dominique Endres, Kimon Runge, Sara Tucci, Anke Schumann, Katharina Domschke, Ludger Tebartz van Elst, Simon Maier

## Abstract

**Background:** Previous research suggests potential mitochondrial dysfunction and changes in fatty acid metabolism in a subgroup of individuals with autism spectrum disorder (ASD), indicated by higher lactate, pyruvate levels, and mitochondrial disorder prevalence. This study aimed to further investigate potential mitochondrial dysfunction in ASD by assessing blood metabolite levels linked to mitochondrial metabolism.

**Methods:** Blood levels of creatine kinase (CK), alanine aminotransferase (ALT), aspartate aminotransferase (AST), lactate, pyruvate, free and total carnitine, as well as acylcarnitines were obtained in 73 adults with ASD (47 males, 26 females) and compared with those of 71 neurotypical controls (NTC) (44 males, 27 females). Correlations between blood parameters and psychometric ASD symptom scores were also explored.

**Results:** Elevated ALT (p = 0.024) and lower CK (p = 0.007) levels were found exclusively in males with ASD compared to NTC, with no such variation in females. AST levels were consistent in both groups. After correction for antipsychotic and antidepressant medication, only CK remained significant. ASD participants had lower serum lactate levels (p = 0.012) compared to NTC, but pyruvate and carnitine concentrations showed no significant difference. ASD subjects had significantly increased levels of certain acylcarnitines, with a decrease in tetradecadienoyl-carnitine (C14:2), and certain acylcarnitines correlated significantly with autistic symptom scores.

**Discussion:** We found reduced serum lactate levels in ASD, in contrast to previous studies suggesting elevated lactate or pyruvate. This difference may reflect the focus of our study on high-functioning adults with ASD, who are likely to have fewer secondary genetic conditions associated with mitochondrial dysfunction. Our findings of significantly altered acylcarnitine levels in ASD support the hypothesis of altered fatty acid metabolism in a subset of ASD patients.

## Introduction

Beyond the core symptoms of stereotypical and repetitive behaviors or interests, individuals with autism spectrum disorder (ASD) often present with increased exhaustibility. This may arise not only from a hypersensitivity to sensory stimuli, but also from the challenges associated with communication, social interaction, and coping with unexpected changes (Lord et al., 2022). ASD prevalence worldwide is estimated at 1-2% (Lord et al., 2022), with prevalence rated in males three to four times higher than in females (Loomes et al., 2017; Maenner, 2020).

The multifactorial underlying etiology of ASD remains largely elusive, with both genetic and environmental factors, such as perinatal injuries, infections during pregnancy, and preterm delivery, potentially playing roles (Lord et al., 2018). Currently, diagnosis is reliant on behavioral criteria, as no validated biomarkers have been identified to support the diagnostic process (Barone et al., 2018).

A significant increase in the prevalence of mitochondrial diseases has been detected in individuals with ASD (≈ 5%) compared to the general population (≈ 0.01%), as revealed in a systematic review and meta-analysis (Rossignol and Frye, 2012). Studies report that between 30-80% of children with ASD show some degree of mitochondrial dysfunction (Frye et al., 2019; Rose et al., 2018). Therefore, it has been proposed that alterations in mitochondrial energy metabolism might serve as potential underlying pathophysiological correlate in a subgroup of individuals with ASD (Weissman et al., 2008). The investigation of mitochondria, crucial cell organelles responsible for energy production and the metabolism of carbohydrates and fats to produce ATP, has been gaining increasing interest in psychiatric research (Kępka et al., 2021).

Previous studies mainly focused on children with ASD, analyzing both direct markers of mitochondrial dysfunction, such as lactate, pyruvate, and acylcarnitine profiles, and indirect markers, including creatine kinase (CK), alanine-aminotransferase (ALT), aspartate-aminotransferase (AST), as well as free and total L-carnitine (Kępka et al., 2021).

Previous studies consistently reported elevated lactate (Al-Mosalem et al., 2009; Hassan et al., 2019; Karim et al., 2016; Oh et al., 2020; Oliveira et al., 2005; Shahjadi et al., 2017) and pyruvate levels (Giulivi et al., 2010; Hassan et al., 2019; Oh et al., 2020) in children with ASD compared to neurotypical controls (NTC). A meta-analysis estimated that 31.1% of children with ASD demonstrated elevated lactate levels, 13.6% had increased pyruvate levels, and 27.6% had an elevated lactate-to-pyruvate ratio (Rossignol and Frye, 2012). However, it is important to note that pyruvate levels and the lactate-to-pyruvate ratio are no longer commonly used in routine diagnostics for mitochondrial disorders. Additionally, not all mitochondrial disorders manifest with elevated lactate levels, and lactate is not specific for mitochondrial dysfunction, as its levels can be influenced by other factors such as physical exercise, hypoxia, and certain medications (Karlsson and Saltin, 1970; Kraut and Madias, 2014).

With regard to blood markers such as CK (Al-Mosalem et al., 2009; Hassan et al., 2019; Khemakhem et al., 2017; Shahjadi et al., 2017), AST (Karim et al., 2016; Poling et al., 2006; Shahjadi et al., 2017), and ALT (Karim et al., 2016), increased activity has been reported. However, given the limited number of studies, further research is required.

Carnitines as amino acid derivates play a key role for mitochondrial metabolism (Malaguarnera and Cauli, 2019). L-carnitine is involved in the transfer of long-chain fatty acids from the cytoplasm to the mitochondria for ß-oxidation, but also stimulates the synthesis of acetylcholine and expression of growth-associated protein-43, along with bolstering neurotransmission and protecting against cell apoptosis and neuron damage (Kępka et al., 2021). Studies in children with ASD have reported significantly reduced levels of both free and total carnitine (Filipek et al., 2004). This finding was further confirmed in a subgroup of individuals with ASD presenting with gastrointestinal abnormalities (Mostafa and Al-Ayadhi, 2015). Clinical trials align with these findings, suggesting that carnitine supplementation may ameliorate autistic symptoms in non-syndromic ASD (Malaguarnera and Cauli, 2019). Consistently, studies found that reduced carnitine levels resulting in a disturbed ß-oxidation of long-chain fatty acids in the fetal nervous stem cells were associated with an increased risk of developing ASD (Bankaitis and Xie, 2019).

In respect to acylcarnitines, a retrospective study that analyzed the acylcarnitine panels of 213 ASD individuals found abnormalities in 17% of children and detected a unique pattern of elevated short and long, but not medium chain acylcarnitines (Frye et al., 2013). Additionally, a prospective study demonstrated that the analysis of acylcarnitines and amino acids from dried blood spots could differentiate ASD and NTC with 73% accuracy in children aged 5 years or younger (Barone et al., 2018).

To elaborate, primary mitochondrial diseases are characterized by defects in genes integral to mitochondrial function or oxidative phosphorylation, while secondary mitochondrial dysfunction arises from diverse genetic and environmental factors not directly related to oxidative phosphorylation. The impairment in secondary mitochondrial dysfunction is often less severe and may not meet the criteria for classification as a primary mitochondrial disease (Niyazov et al., 2016). The prevalence of altered biomarkers of mitochondrial dysfunction was reported to be higher than those of mitochondrial disease and most cases were not associated with genetic abnormalities which points toward a possible secondary mitochondrial dysfunction (Rossignol and Frye, 2012). In a review, Citrigno et al. (2020) come to the conclusion that the relationship between specific genetic variants and their resulting traits in autism spectrum disorder (ASD) is still unclear and that it remains difficult to determine if mitochondrial dysfunctions are a cause or result of ASD.

### Aims of the Study

This study’s objective was to investigate potential markers of mitochondrial dysfunction in adults with ASD, based on the hypothesis of a higher incidence of mitochondrial anomalies in these individuals compared to NTC. We analyzed both direct markers of mitochondrial dysfunction, such as lactate, pyruvate, and acylcarnitine profiles, and indirect parameters, which include CK, ALT, AST, free and total L-carnitine. Based on prior studies, we hypothesized to find elevated lactate, pyruvate, and lactate-to-pyruvate ratios, along with increased CK, AST, and ALT levels. With regard to carnitine and free carnitine levels, we expected decreased levels and a distinct acylcarnitine profile characterized by reduced levels of short and long-chain acylcarnitines.

## Methods

### Participants

The study was conducted at the Department of Psychiatry and Psychotherapy of the University of Freiburg, Germany, in accordance with the Declaration of Helsinki, following written informed consent from all participants. The Ethics Committee of the University of Freiburg, Germany, approved the study (Approval ID: 268/17).

Patients with the diagnosis of ASD according to the Diagnostic and Statistical Manual of Mental Disorders (DSM-5; 299.00) and Asperger-Syndrome according to the International Classification of Diseases (ICD-10; F84.5) were enrolled in the study. The diagnoses were determined by experienced senior psychiatrists. To obtain a pathomechanistically homogeneous sample, we specifically excluded individuals with recognized secondary genetic forms of autism spectrum disorders (ASD). The NTC group was recruited via advertisements.

All study participants were required to complete the Autism Spectrum Quotient (AQ) (Baron-Cohen et al., 2001), the Empathy Quotient (EQ) (Baron-Cohen and Wheelwright, 2004), as well as the Social Responsiveness Scale 2 (SRS-2) (Constantino and Gruber, 2012) to assess autistic symptoms in the ASD group and rule them out in the NTC group. Moreover, the Beck Depression Inventory (BDI-II) (Hautzinger et al., 2006) was used to assess comorbid depressive symptoms, the Conners’ Adult ADHD Rating Scales (CAARS) (Conners et al., 1999) and the Wender Utah Rating Scale (WURS-k) (Retz-Junginger et al., 2002) were used to evaluate symptoms of attention-deficit/hyperactivity (ADHD) disorder. Crystallized intelligence (IQ) was estimated with the Multiple Choice Vocabulary Test (MWT-B) (Lehrl, 2005). To assess the impairment due to physical and in particularly mental symptoms, the Symptom-Checklist-90^®^ (SCL-90-R) was used (Derogatis and Savitz, 1999). After the blood sampling and before entering the scanner bore, we assessed the subjects state of anxiety using the state anxiety inventory (Spielberger et al., 1970).

Exclusion criteria were defined for all study participants, encompassing general contraindications for magnetic resonance imaging (MRI) (e.g. pregnancy, metallic implants, claustrophobia), diabetes mellitus, obesity (BMI > 30 kg/m^2^), neurological disorders (e.g., seizures), the intake of benzodiazepines). For ASD participants, additional exclusion criteria were bipolar disorders, psychotic symptoms, and substance abuse. Only participants devoid of psychiatric disorders were included in the NTC group.

The sample was also part of a magnetic resonance tomography study focusing on lactate spectroscopy, of which the results will be published separately.

### Sample collection and analysis

Serum, ethylenediaminetetraacetic acid (EDTA) treated blood, and dried blood spot cards were utilized for various analyses for both ASD patients and NTC. CK, ALT, and AST were assessed in the serum, whereas pyruvate, lactate, and the lactate-to-pyruvate ratio were examined in EDTA-treated blood. Dried Blood Spot Filter Paper cards were prepared for the analysis of free and total carnitine as well as acylcarnitines. The Institute for Clinical Chemistry and Laboratory Medicine, Medical Center, University of Freiburg performed the assays for CK, ALT, and AST. The Center for Pediatrics and Adolescent Medicine, Medical Center, University of Freiburg analyzed lactate, pyruvate, carnitine, and acylcarnitine levels.

#### CK, ALT, and AST

CK levels were assessed using the Roche/Hitachi cobas® c 701 system, an automated photometric analyzer. The method was adapted from the protocol of the International Federation of Clinical Chemistry (IFCC) and optimized for performance and stability. CK catalyzes the conversion of creatine phosphate and adenosine diphosphate (ADP) into creatine and Adenosine triphosphate (ATP). Then, hexokinase catalyzes the reaction of ATP and D-glucose to form ADP and Glucose-6-phosphate (G6P). Subsequently, glucose-6-phosphate dehydrogenase (G6PD) catalyzes the transformation of G6P, along with Nicotinamide adenine dinucleotide phosphate (NADP^+^), into D-6-phosphogluconate, NADPH, and H^+^. The photometrically determined rate of NADPH formation is directly proportional to the CK activity.

ALT and AST were also analyzed with the Roche/Hitachi cobas® c system. The test is based on an optimized version of the IFCC recommendations. The addition of pyridoxal phosphate to the assay increases the aminotransferase activity and avoids an underestimation of the aminotransferase activity in samples with insufficient endogenous pyridoxal phosphate. The activation for AST is higher than for ALT.

ALT catalyzes the conversion of L-alanine and 2-oxoglutarate into pyruvate and L-glutamate. The generated pyruvate is subsequently reduced to L-lactate and NAD^+^ by NADH, in a reaction catalyzed by Lactate Dehydrogenase (LDH). The oxidation rate of NADH, which is proportional to the catalytic activity of ALT, is measured by determining the extinction decrease.

AST catalyzes the transfer of an amino group between L-aspartate and 2-oxoglutarate to form oxalacetate and L-glutamate. In the presence of malate dehydrogenase (MDH), oxalacetate then reacts with NADH to generate NAD+. Pyridoxal phosphate serves as a coenzyme in the aminotransfer reaction, ensuring full enzyme activation. The rate of NADH oxidation, which is directly proportional to the catalytic activity of AST, is measured by determining the extinction decrease.

The following upper threshold reference values were defined: CK (male) = 190 U/L, CK (female) = 170 U/L, ALT (male) = 50 U/L, ALT (female) = 35 U/L, AST (male) = 50 U/L, and AST (female) = 35 U/L.

#### Lactate, pyruvate, and lactate-to-pyruvate-ratio

Lactate levels were analyzed with the “Lactate Assay Kit MAK064”, while the “Pyruvate Assay Kit MAK071” by Sigma-Aldrich® was used for pyruvate analysis.

With the “Lactate Assay Kit MAK064”, the lactate concentration is determined by an enzymatic assay that leads to a colorimetric (570 nm)/fluorometric (□_ex_ = 535 nm/□_em_□ = 587 nm) product, proportional to the lactate concentration. Typical detection ranges for this kit are between 0.2-10 nmoles of lactate.

In the case of the “Pyruvate Assay Kit MAK071”, the pyruvate concentration is obtained by a coupled enzyme assay that results in a colorimetric (570 nm)/fluorometric (λ_ex_ = 535/λ_em_ = 587 nm) product, proportional to the pyruvate concentration.

#### Free and total carnitines, acylcarnitines

Acylcarnitines were extracted from the dried blood spots (corresponding to 50µL of blood) using a solution of acetonitrile/water (80/20% v/v) that contained isotopically labeled acylcarnitines of various chain lengths at predetermined concentrations as internal standard. Afterwards, the extract was dried under nitrogen and derivatized by the addition of n-butanol HCl to yield the acylcarnitines for analysis. The acylcarnitines were measured as their n-butyl-esters and quantitative concentrations of the analytes were calculated by comparison of the detected abundance of each acylcarnitine against that of a designated internal standard with a known concentration (Smith and Matern, 2010).

The internal standard applied was MassChrom Amino Acids and Acylcarnitines from dried blood by Chromsystems®, Gräfeling, Germany. The normative range for free and total carnitine, as well as all other acylcarnitines, was provided by the processing laboratory.

Free carnitine and acylcarnitine levels were measured using electrospray tandem mass spectrometry (ESI/MS/MS) based on a highly sensitive and specific method of Vreken et al. (1999). This gentle ionization technique has the advantage to minimize molecule fragmentation (Vreken et al., 1999). In this study, for the majority of acylcarnitine profiles (124 of 143), this system was used. However, due to an equipment upgrade, the last 19 of the 143 acylcarnitine profiles were measured with a newer ESI/MS/MS system.

The concentrations of total and free carnitine as well as the following acylcarnitines were measured: free carnitine/palmitoyl-carnitine (C0/C16), acetyl-carnitine (C2), propionyl-carnitine (C3), propionyl-carnitine/palmitoyl-carnitine (C3/C16), butyryl-carnitine (C4), hydroxy-butyryl-carnitine (C4-OH), tiglyl-carnitine (C5:1), isovaleryl-carnitin+3-hydroxy-butyryl-carnitine (C5+C4:1-OH), 3-hydroxy-isovaleryl-carnitine (C5-OH), hexanoyl-carnitine (C6), hexenoylcarnitine (C6:1), 3-hydroxy-hexanoyl-carnitine (C6-OH), octanoyl-carnitine (C8), octanoyl-carnitine/decanoyl-carnitine (C8/C10), octanoyl-carnitine/lauroyl-carnitine (C8/C12), octanoyl-carnitine/acetylcarnitine (C8/C2), decanoyl-carnitine (C10:1), decadienoyl-carnitine (C10:2), 3-hydroxy-dodecanoylcarnitine (C12-OH), tetra-decanoyl-carnitine (C14:1), tetradecadienoyl-carnitine (C14:2), octadecenoyl-carnitine (C18:1), 3-hydroxy-oleyl-carnitine (C18:1-OH), octadecadienyl-carnitine (C18:2), 3-hydroxy-linoleyl-carnitine (C18:2-OH).

### Statistical analysis

Statistical analyses were performed using “R” (R Core Team, 2022) within the Rstudio environment (RStudio Team, 2022).

In cases where two-sample t-tests were applied, data normality was verified using the Shapiro-Wilk test. For all linear models, normality of residuals was checked with the Shapiro-Wilk test, and homogeneity of variances was assessed via the Levene-Test.

#### Psychometric parameters

To compare psychometric data across groups, two-sample t-tests were employed for age, IQ, AQ, EQ, SRS-2, WURS-k, and BDI-II, alongside a Pearson’s Chi-squared test for sex.

#### Blood data and dried blood spot cards

For data violating the assumption of normality, a log-transformation was performed. A number of blood parameters remained non-normally distributed even after log-transformation, necessitating the application of Tukey’s Ladder of Powers transformation for all data violating the assumption of normality (# https://rcompanion.org/handbook/I_12.html; retrieval date: 17^th^ May, 2023)

#### CK, ALT, and AST

Group differences in elevated CK (upper threshold male = 190 U/L, female = 170 U/L), ALT (upper threshold male = 50 U/L, female = 35 U/L), and AST (upper threshold male = 50 U/L, female = 35 U/L) were examined via Fisher’s Exact Test.

CK, ALT, and AST were analysed separately for female and male participants, and these variables were subjected to a Tukey’s Ladder of Powers transformation to meet the requirement of normal distribution. Group differences in mean CK, ALT, and AST were evaluated applying two-sample t-tests. All six t-tests were subsequently corrected for multiple comparisons applying the Benjamini Hochberg approach (Benjamini and Hochberg, 1995).

The influence of age on CK, ALT, and AST levels was examined applying separate linear models for females and males.

#### Lactate & Pyruvate

Group differences in elevated lactate (upper threshold = 2 mmol/L), pyruvate (upper threshold = 130 μMol/L), and lactate-to-pyruvate ratio (upper threshold = 17) were examined using Fisher’s Exact Test.

While pyruvate was normally distributed, lactate maintained a significant skewness even after log-transformation, necessitating a Tukey’s Ladder of Powers transformation. Despite an improvement in the skewness of the lactate concentrations, the requirement for normal distribution was not met.

Group differences in mean Tukey-transformed lactate levels were assessed applying a Yuen’s independent samples t-test for trimmed means (WRS2 package) (Wilcox, 2017; Yuen, 1974). All three t-tests were then adjusted for multiple comparisons applying the Benjamini-Hochberg procedure.

To test for the influence of age and sex on lactate and pyruvate levels, a linear (pyruvate) and a robust linear model (lactate; lmrob package) were utilized.

#### Carnitine & Acylcarnitine

Group differences in reduced levels of free carnitine (lower threshold 15 μmol/L) and total carnitine (lower threshold = 20 μmol/L) were assessed using Fisher’s Exact Test.

Neither free nor total carnitine were normally distributed according to the Shapiro-Wilk normality test, which necessitated log transformation of the data. A linear model was used for group effects, which was corrected for potential confounding effects of age, sex, and the type of NMR-spectrometer system. For plotting, the concentration was adjusted for the influence of NMR-spectrometer and sex.

Acylcarnitines also deviated from a normal distribution and were therefore log-transformed. A linear model was then used to test for potential confounding effects of age, sex, and the type of NMR-spectrometer system. Since age and sex displayed only minor confounding effects on a very limited number of acylcarnitines, the log-transformed concentration was only adjusted for the effect of type of NMR-spectrometer system using the predict function of the R stats package. The group differences of the log-transformed and adjusted acylcarnitines concentration between ASD and NTC were then compared using two-sample t-tests. Both unadjusted p-values and p-values adjusted for multiple comparisons are reported.

#### Correlations of psychometric scores and blood parameters

Spearmen’s correlations were calculated between psychometric scores (AQ (Baron-Cohen et al., 2001), EQ (Baron-Cohen and Wheelwright, 2004), SRS-2 (Constantino and Gruber, 2012) total scores, depressive symptoms according to the BDI-II (Hautzinger et al., 2006), IQ assessed with the MWT-B (Lehrl, 2005) and the adjusted blood parameters (CK, ALT, AST, total and free carnitine, acylcarnitines). If required, blood parameters were adjusted for confounding effects of sex (CK, ALT, AST) or the NMR-spectrometer (total and free carnitines, acylcarnitines). Adjusted p-values for multiple comparisons according to the Benjamini Hochberg approach (Benjamini and Hochberg, 1995) are reported.

#### Potential influence of medication

We conducted a secondary analysis by re-running all statistical models that exhibited significant group effects or differences, this time accounting for the potential influence of anti-psychotic and anti-depressive medications, as well as the intake of oral contraceptives, to ascertain the robustness of our initial findings.

## Results

### Participants

The study comprised 73 adults diagnosed with ASD (47 male, 26 female) and 71 NTC (44 male, 27 female). Both groups showed no significant difference with regard to IQ as assessed with the MWT-B (Lehrl, 2005). As anticipated, the ASD group showed elevated scores in AQ (Baron-Cohen et al., 2001) and SRS-2 (Constantino and Gruber, 2012), whereas the EQ (Baron-Cohen and Wheelwright, 2004) was higher in the NTC group. Participants with ASD exhibited significantly higher levels of depressive symptoms, as measured by the BDI-II (Hautzinger et al., 2006), and showed greater anxiety as indicated by the STAI. Additionally, they exhibited more symptoms indicative of ADHD in childhood, when compared to NTC. Only patients were prescribed psychiatric medications, leading to significantly higher usage of typical and atypical antipsychotics, SNRIs, SSRIs, and other antidepressants. The difference in the intake of Lithium, Stimulants, Anticonvulsants, standby Benzodiazepines, and Oral contraceptives between groups wasn’t significant. Table 1 summarizes psychometric parameters of both the ASD and NTC group.

**Table 1:** Demographic and psychometric data.

| Parameter |  | ASD (N = 73) | NTC (N = 71) | p-value |
| --- | --- | --- | --- | --- |
| Sex (male/female) | Count | 47/26 | 44/27 | .764 |
| Age in years | Mean<br>(SD) | 36.2 (12) | 31.7 (9.8) | .015 |
| IQ (MWT-B) |  | 115 (14.3) | 114.5 (13.9) | .841 |
| SRS-2 |  | 108.2 (28.8) | 34.9 (14.5) | < .001 |
| AQ |  | 35.6 (8.1) | 11.8 (5) | < .001 |
| EQ |  | 21.8 (11.2) | 47.4 (10.4) | < .001 |
| BDI-II |  | 15.7 (13.1) | 3.8 (4.5) | < .001 |
| WURS-k |  | 27.7 (15.8) | 13.0 (10.3) | < .001 |
| STAI |  | 24.5 (10.8) | 11.6 (6.4) | < .001 |
| Typical antipsychotics | Count | 8 (11%) | 0 (0%) | 0.006 |
| Atypical antipsychotics |  | 7 (9.6%) | 0 (0%) | 0.013 |
| SNRIs / SSRIs |  | 16 (22%) | 0 (0%) | <0.001 |
| Other antidepressants |  | 16 (22%) | 0 (0%) | <0.001 |
| Lithium |  | 2 (2.7%) | 0 (0%) | 0.5 |
| Stimulants |  | 4 (5.5%) | 0 (0%) | 0.12 |
| Anticonvulsants |  | 1 (1.4%) | 0 (0%) | >0.9 |
| Benzodiazepines (standby) |  | 2 (2.7%) | 0 (0%) | 0.5 |
| Oral contraceptives |  | 3 (4.1%) | 5 (7.0%) | 0.5 |

Abbreviations: AQ = Autism Spectrum Quotient (Baron-Cohen et al., 2001); BDI-II = Beck Depression Inventory II (Hautzinger et al., 2006); EQ = Empathy Quotient (Baron-Cohen and Wheelwright, 2004); IQ = Intelligence Quotient; MWT-B = Multiple Choice Vocabulary Test (Lehrl et al., 1995); SRS-2 = Social Responsiveness Scale 2 (Constantino, 2013); WURS-k = Wender Utah Rating Scale (Retz-Junginger et al., 2002); STAI = State Trait Anxiety Inventory (Spielberger et al., 1970); SNRI = serotonin-norepinephrine reuptake inhibitor; SSRI = selective serotonin reuptake inhibitor.

### CK, ALT, and AST

No significant differences in CK (p = 0.972, p_corr_ = 0.972), ALT (p = 0.331, p_corr_ = 0.662), and AST (p = 0.824, p_corr_ = 0.972) were detected between female individuals with ASD and NTC. However, male ASD participants showed lower CK (p = 0.007, p_corr_ = 0.045) and ALT (p = 0.024, p_corr_ = 0.073) levels compared to NTC. Age did not significantly effect on CK (female: p=0.179; male: p=0.119), ALT (female: p=0.636; male: p=0.489), and AST (female: p=0.270; male: p=0.131) concentrations. Figure 1 depicts CK, ALT, and AST levels in female and male individuals with ASD compared to NTC. In the secondary analysis, group effects in CK between male participants with ASD and NTC persisted when correcting for the influence of antipsychotic (p = 0.022) and antidepressant (p = 0.011) medication. However, for ALT, the p-value for the group effect in male participants increased to 0.084 when correcting for anti-depressants and 0.062 when correcting for anti-psychotic medication, indicating no statistical significance.

**Figure 1:**
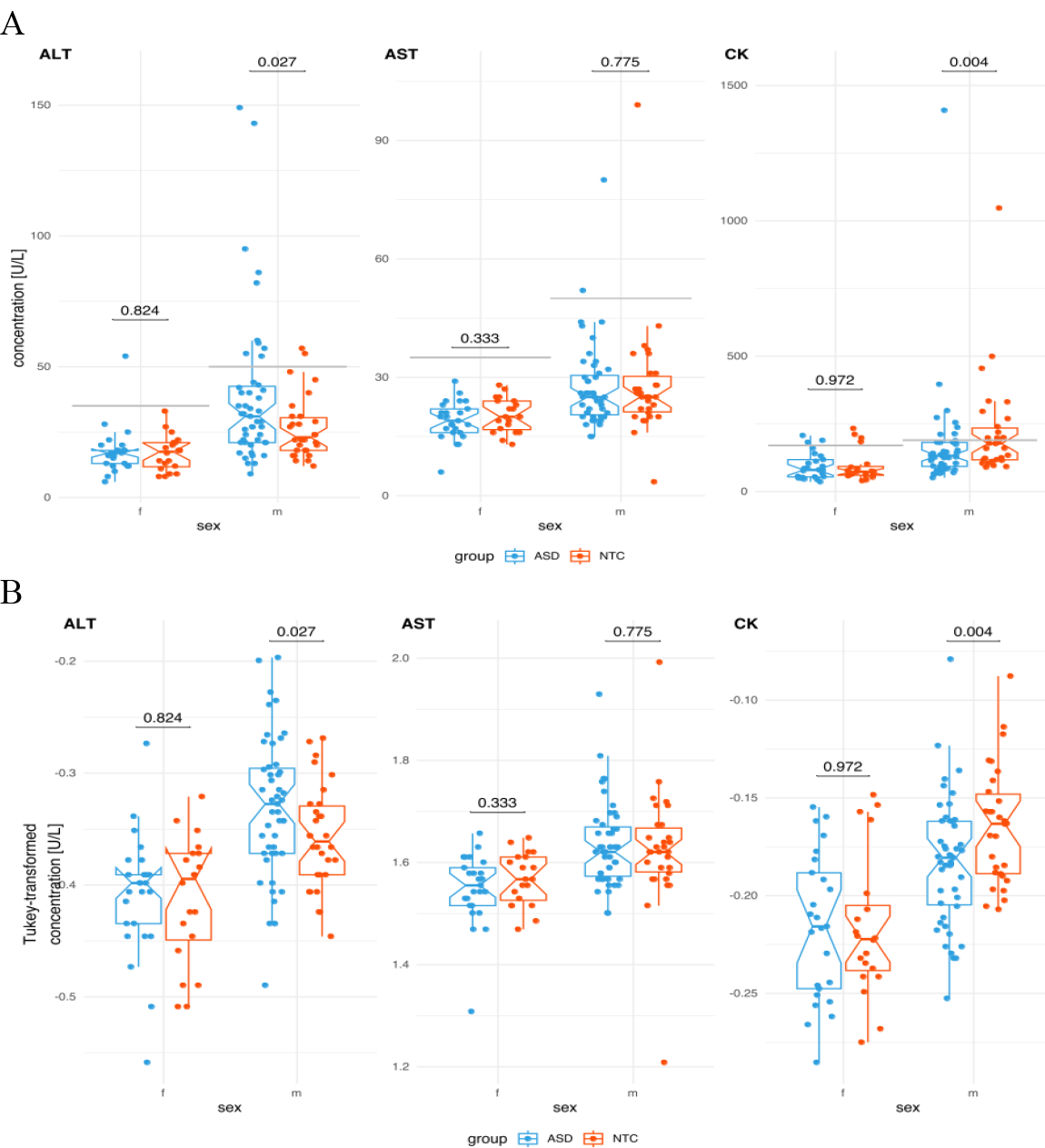
Box-plots of AST, ALT, and CK levels in individuals with ASD compared to NTC separately for female and male participants. A: Raw concentrations B: Tukey-transformed concentrations. Male participants of the ASD group exhibited elevated ALT and lower CK levels. Notches depict ± one standard deviation. Grey horizontal lines shows the according reference value. Black horizontal lines with numbers show the p-value of the according group comparison.

### Lactate, pyruvate, and lactate-to-pyruvate ratio

Individuals with ASD showed lower serum lactate levels in comparison to NTC (p = 0.012, p_corr_ = 0.036) (Figure 2), while no significant difference in pyruvate concentrations (p=0.258, p_corr_ = 0.387) or the lactate-to-pyruvate ratio (p = 0.544, p_corr_ = 0.544) was detected between ASD and NTC groups. Both age and sex had no significant impact on lactate (age: p=0.113; sex: p=0.253) and pyruvate (age: p=0.750; sex: p=0.567) concentrations. The group effects for serum lactate levels remained significant when incorporating antidepressants (p = 0.0249), antipsychotics (p = 0.003), and oral contraceptives (p = 0.018) as covariates.

**Figure 2:**
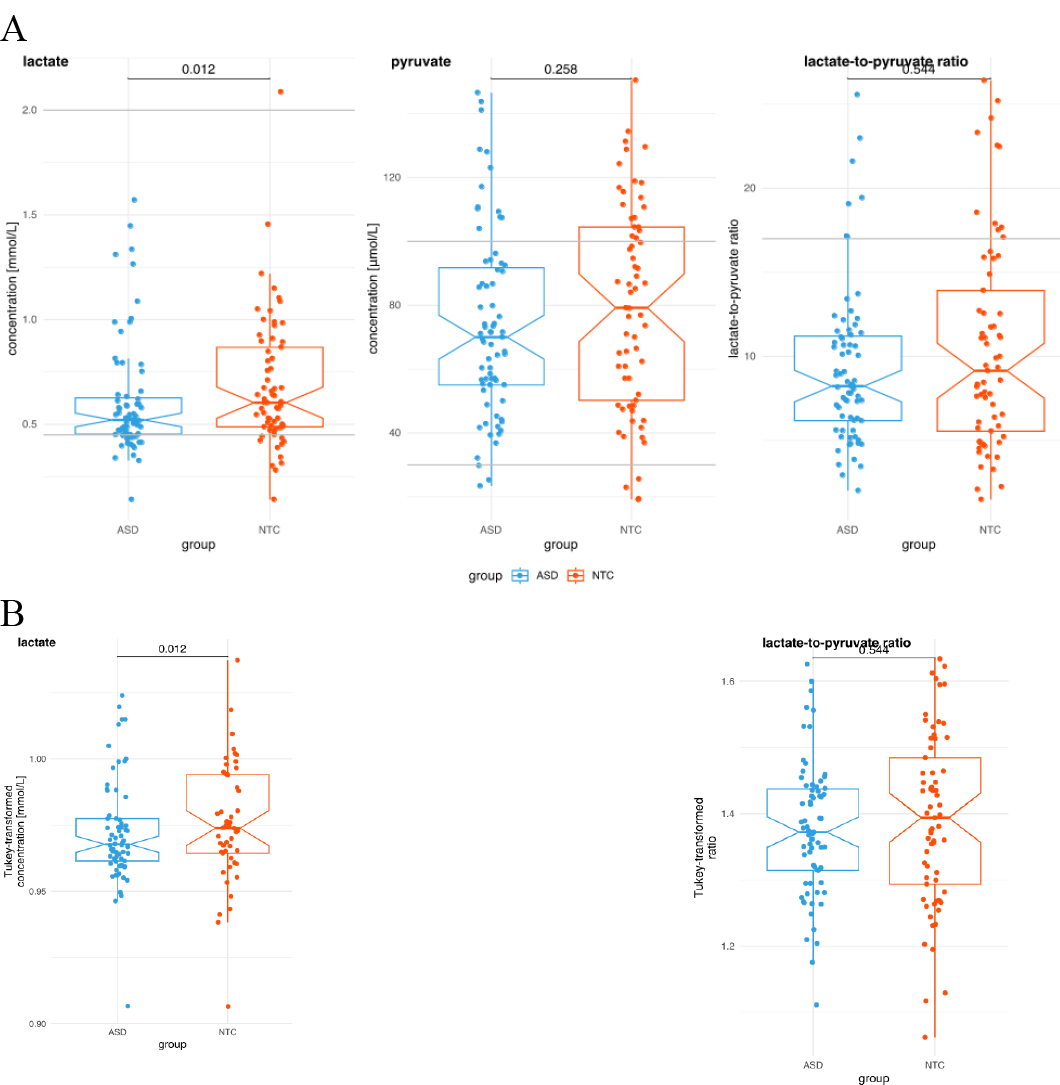
Box-plots of lactate, pyruvate and lactate-to-pyruvate ratio in individuals with ASD compared to NTC. A: Raw concentrations B: Tukey-transformed concentrations for lactate and lactate-to-pyruvate ratio (pyruvate was normally distributed). Participants with ASD exhibited lower serum lactate levels. Notches depict standard deviation. Grey horizontal lines shows the according reference value. Black horizontal lines with numbers show the p-value of the according group comparison.

### Total and free carnitine

No significant influence was observed for the factors group (total carnitine: p=0.391; free carnitine: p=0.242) and age (total carnitine: p=0.083; free carnitine: p=0.186) on total and free carnitine levels. However, both sex (total carnitine: p=0.001; free carnitine: p<0.001) and NMR-spectrometer (total carnitine: p<0.001; free carnitine: p<0.001) had significant effects. Figure 3 shows the box-plots of total and free carnitine levels in individuals with ASD compared to NTC.

**Figure 3:**
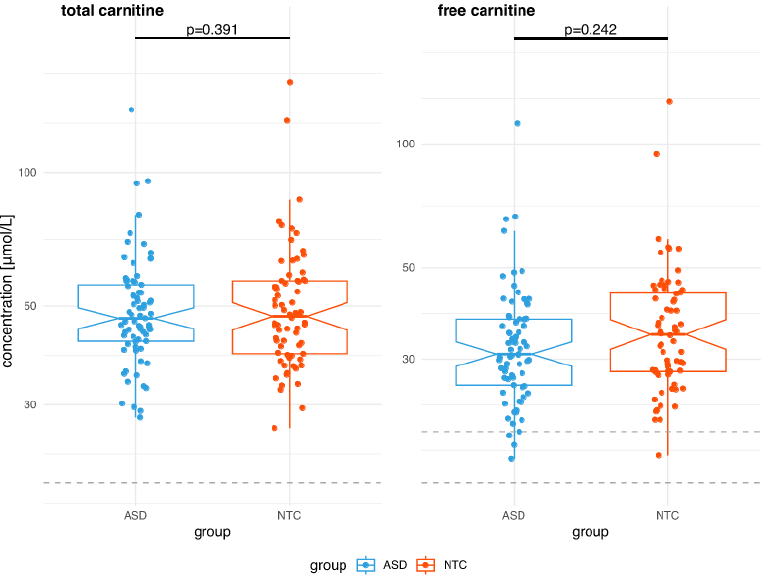
Box-plots of total and free carnitine levels in individuals with ASD compared to NTC. Adjusted concentrations (sex and NMR-spectrometer) for total and free carnitine [y-axis log-transformed]. Notches depict standard deviation. Grey horizontal lines shows the according lower reference value. Black horizontal lines with numbers show the p-value of the group effect of the according linear model described in the methods section. No group differences in total and free carnitine between ASD and NTC.

### Acylcarnitines

Following adjustment for multiple comparisons, individuals with ASD exhibited elevated levels of several log-transformed and NMR-spectrometer-adjusted acylcarnitines (C5:1, C6, C6:1, C6-OH, C10:1, C10:2) in comparison to NTC, with the exception of C14:2, which was diminished in the ASD group (Figure 4). Notably, upon controlling for the impact of anti-psychotic medication, the group effects remained significant for the aforementioned acyl-carnitines, with the addition of C18:1-OH. Similarly, after accounting for antidepressants, the group effects persisted for all previously mentioned acylcarnitines, except for C10:1, which did not attain significance, while C18:1-OH emerged as significant. After adjusting for the intake of oral contraceptives, the group effects persisted for all previously mentioned acylcarnitines, while additionally C8:C12 and C18:1-OH emerged as significant.

**Figure 4:**
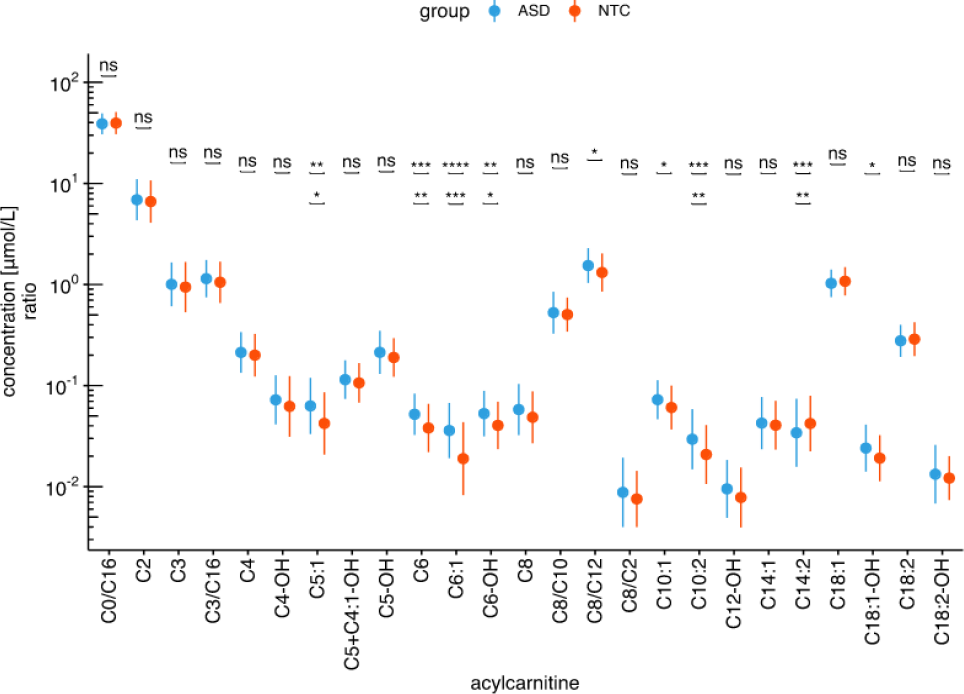
Acylcarnitine levels in individuals with ASD compared to NTC. Adjusted concentrations (NMR-spectrometer) for acylcarnitines [y-axis log-transformed]. Notches depict standard deviation. Black horizontal lines with asterisk indicate the significance level of the t-test’s group differences before and after adjustment for multiple comparisons [lower line]. ns=not significant; * = p<.05; ** = p<.01; *** = p<.001; **** = p<.0001

### Correlations

Significant positive correlations were observed between the adjusted acylcarnitine concentrations of C5:1, C6, C6:1, C6-OH, and C10:2, and the total scores of the autism spectrum quotient (AQ) (Baron-Cohen et al., 2001) and the SRS-2 (Constantino and Gruber, 2012). The acylcarnitines C5:1, C6, C6:1, C6-OH, and C10:2 were negatively correlated with the empathy quotient (EQ total) (Baron-Cohen and Wheelwright, 2004). In addition, C6 and C6:1 displayed a positive associated with the Beck Depression Inventory (BDI-II) (Hautzinger et al., 2006) (Figure 5).

**Figure 5:**
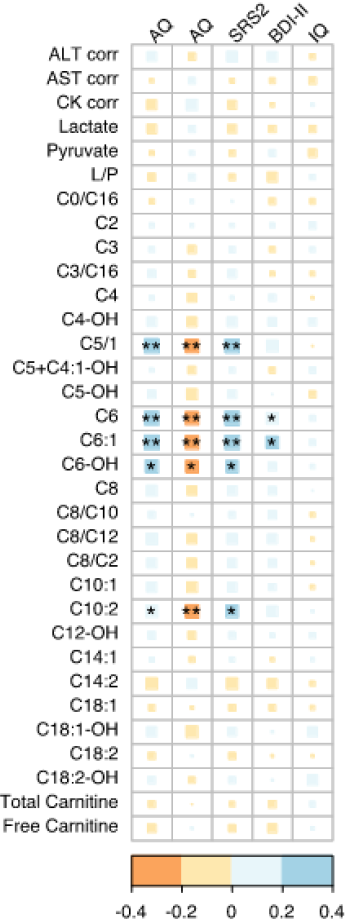
Correlation matrix of psychometric scores and blood parameters in individuals with ASD and NTC across both groups. Psychometric scores (AQ, EQ and SRS-2 total score, BDI-II, IQ assessed with the MWT-B) were correlated with blood parameters (adjusted for confounding factors if necessary: CK (sex), ALT (sex), AST (sex), total and free carnitines (NMR-spectrometer), acylcarnitines (NMR-spectrometer)). Results adjusted for multiple comparison according to Benjamini-Hochberg. *p < 0.05 **p < 0.01

## Discussion

In this study, we analyzed a variety of different markers of mitochondrial metabolism in adults with ASD as compared to NTC. Lower CK and elevated ALT levels were detected in male ASD patients relative to NTC while no alterations for these were found in female individuals with ASD nor for AST in both genders. However, after adjustment for medication, only CK values remained significantly different. Lactate levels were lower in the ASD group, while no significant differences in pyruvate, lactate-to-pyruvate ratio, or total and free carnitine levels could be observed. We also detected a specific acylcarnitine profile in the ASD group, characterized by an increase in various acylcarnitines, with the exception of C14:2 which was reduced relative to NTC.

In contrast to previous findings who reported increased levels of serum lactate (Al-Mosalem et al., 2009; El-Ansary, 2010; Hassan et al., 2019; Karim et al., 2016; Mostafa et al., 2005; Oh et al., 2020; Shahjadi et al., 2017) and pyruvate (Giulivi et al., 2010; Hassan et al., 2019; Oh et al., 2020), we found lower lactate and unaltered pyruvate levels in ASD compared to NTC. While laboratory assays might contribute to variance in measurements, they are unlikely to be the sole reason for the discrepancy in lactate levels observed in our study. Although increased levels of state anxiety have been associated with elevated lactate levels (Maddock et al., 1991), this does not seem to be the driving factor here as we observed higher state anxiety levels in our ASD group but, contrary to expectations, found decreased lactate levels. Additionally, the procedure for blood sample collection may influence lactate levels; anxious children may take longer to cooperate during blood drawing, which could prolong the tourniquet application and possibly lead to a transient increase in lactate levels due to local hypoxia (Benzon et al., 1988). Lastly, our study differed from previous research in terms of the sample characteristics. Previous studies mainly involved children with ASD and did not exclude cases with secondary genetic conditions that may affect mitochondrial function. In contrast, our study focused on high-functioning adults, who might be less affected by secondary genetic conditions influencing mitochondrial function and metabolism. Consequently, the decreased lactate levels observed in our study, which are seemingly at odds with prior findings, might be indicative of a different metabolic profile specific to the high-functioning adult ASD subgroup under investigation

In our study, there were no significant differences in the levels of CK, ALT, and AST between female adults with and without ASD. Conversely, male adults with ASD exhibited lower CK and elevated ALT levels compared to NTC. However, ALT levels, only showed a significant difference before adjustment for medication. Although all participants were asked to refrain from exercise the day before the measurements, it is worth noting that CK levels can remain elevated for a longer period after exercise (Brancaccio at al., 2007). It is possible that the NTC group in general participated more frequently in sports or other physical activities, leading to a relative increase in their CK levels.

Still, this observation raises questions about possible sex differences in the expression of these enzymes in ASD. Previous studies in this area reported increased CK levels, but they either exclusively included male participants or have not differentiated their findings based on sex (Al-Mosalem et al., 2009; Hassan et al., 2019; Khemakhem et al., 2017; Shahjadi et al., 2017). This lack of differentiation may have obscured potentially sex-specific variations in CK levels within the ASD population. In contrast to these prior studies, we found no significant differences in AST levels. Elevated serum levels of these enzymes (CK, ALT, AST) may be indicative of cellular integrity loss in tissues such as muscle and liver, a consequence often associated with mitochondrial dysfunction (Hassan et al., 2019).

In contrast to previous studies reporting reduced serum carnitine levels in ASD children and those presenting with gastrointestinal manifestations (Filipek et al., 2004; Mostafa and Al-Ayadhi, 2015), we detected no alterations. Carnitine is an essential nutrient that plays a pivotal role in energy metabolism by transporting long-chain fatty acids into mitochondria for oxidation and subsequent energy production (Virmani and Cirulli, 2022). In the context of our study, it is important to consider several factors that might explain the lack of observed differences in free carnitine levels among adults. Firstly, the heterogeneity of ASD may result in distinct metabolic profiles across the spectrum, especially considering our focus on high-functioning ASD adults who might exhibit different metabolic characteristics than those with more severe forms of ASD. Secondly, dietary and lifestyle factors that can significantly influence carnitine levels are more autonomously controlled in adults than in children, potentially contributing to the similar carnitine levels observed between adult ASD individuals and neurotypical controls in our study.

Our findings suggest a distinctive acylcarnitine profile in individuals with ASD compared to NTC, a potential indicator of impaired mitochondrial fatty acid ß-oxidation. Consistently with our findings of increased acylcarnitine levels, children diagnosed with ASD (Barone et al., 2018) exhibited significantly increased levels of several different acylcarnitines. Interestingly, levels of acetylcarnitine C14:2 are elevated in children with ASD but diminished in our adult sample with ASD, potentially indicating a shift in metabolic status as patients with ASD age. In contrast, in patients with schizophrenia mainly decreased acylcarnitine levels have been reported (Cao et al., 2019). The differing acylcarnitine profiles found in ASD and schizophrenia may provide an opportunity to discern potential metabolic biosignatures. Future studies have to validate and maybe combine so far preliminary biomarkers to support diagnosis of ASD and personalize treatment options.

### Limitations

Altered lactate and CK levels may be a result of physical exercise, blood collection (fist clenching and unclenching) (Vargas et al., 2005) and medication. Participants were not investigated before breakfast and were only requested to not engage in physical activity the day before measurement. Therefore, we cannot exclude a biasing effect due to nutrition or longer lasting effects of sport activities. For example red meat was reported to be a rich source of L-carnitine in adults and milk in infants and children (Kępka et al., 2021).

### Summary

In our study of mitochondrial metabolism in high-functioning adults with ASD vs. NTC, we found male-specific increases in ALT and decreases in CK levels, with only CK levels remaining significantly altered after adjustment for medication. In addition, we found lower lactate levels and an acylcarnitine profile characterized by increases in several acylcarnitines, except for a decrease in C14:2. These findings differ from previous studies in children and may reflect metabolic differences within ASD subgroups. Importantly, our study uniquely accounts for sex-specific variations in CK and ALT levels. No differences were found in AST, total and free carnitine levels, suggesting potential influences of adult-autonomous dietary or lifestyle factors. The different acylcarnitine profiles between ASD and NTC highlight potential metabolic biomarkers. Future research should further validate these findings and explore the integration of these preliminary biomarkers into ASD diagnosis and personalized treatment.

## Data Availability

All data produced in the present study are available upon reasonable request to the authors

## Availability of data and materials

Demographic, psychometric, serum and dried blood spot data as well as the R code for statistical analysis, are available from the corresponding author on request.

## Ethics approval and consent to participate

The Ethics Committee of the University Medical Center Freiburg (Approval ID: 268/17) approved the study. All participants gave written informed consent to participate.

## Authors’ contributions

KN and SM wrote the paper. SM, KN and MM performed the data and statistical analysis. SM and LTvE organized the study and created the study design. ST and AS performed the analysis of lactate, pyruvate and acylcarnitines. KN, DE and LTvE recruited the patients and established the diagnosis. LTvE, MM, DE, ST, AS, KR and KD revised the manuscript critically focusing on clinical and statistical aspects. All authors were critically involved in the theoretical discussion and composition of the manuscript. All authors read and approved the final version of the manuscript.

